# Determinants of healthcare providers’ satisfaction with electronic medical records system (EMRs) in public healthcare facilities in the Ashanti region of Ghana: A multicenter analytical cross-sectional survey

**DOI:** 10.64898/2025.12.03.25341569

**Authors:** Bernard Gyamfi Anane, Victoria Bam, Gertrude Boafo, Emmanuel Osei-Peprah, Jacob Tetteh

**Affiliations:** School of Nursing and Midwifery, College of Health Sciences, Kwame Nkrumah, University of Science and Technology; Effiduase Government Hospital, Ghana Health Service, Ashanti Region

**Keywords:** Lightwave Health Information Management System, determinants, satisfaction, Ashanti region, Ghana

## Abstract

**Background:** Electronic medical record (EMR) systems are increasingly being adopted in low-and middle-income countries to improve healthcare delivery. User satisfaction is crucial for the effective utilisation and long-term sustainability of the system. Despite the scale-up of the Lightwave Health Information Management System (LHIMS) in Ghana, evidence on healthcare providers’ satisfaction remains limited. This study assessed satisfaction with LHIMS and determinants of satisfaction with the EMR among healthcare providers in the Ashanti Region, Ghana.

**Methods:** A multicenter analytical cross-sectional study was conducted from August to October 2024 in the Ashanti Region. Using multistage sampling, 438 healthcare providers were selected from three healthcare facilities with functional LHIMS. Data were collected using a pre-tested, self-administered questionnaire adapted from validated instruments and analysed using IBM SPSS version 25. Descriptive statistics were used to summarise the data, with categorical variables presented as frequencies and percentages, and continuous variables as means with standard deviations, or medians with interquartile ranges, where appropriate. Logistic regression analysis was performed to identify determinants of satisfaction, with results reported at a 5% significance level using adjusted odds ratios and 95% confidence intervals.

**Results:** The mean age of the participants was 33 years (*SD* = 5.7), and 66% were female. Most participants (94%) had received LHIMS training, half (50%) reported adequate access to computers, and 53% had low eHealth literacy. Regarding perceived system qualities, 57.5% rated usability as good, and 50.3% rated system quality as good. Overall, 57% (95% CI: 52–62%) of participants were satisfied with LHIMS.

Multivariable analysis showed that satisfaction was independently associated with good perceived information quality (AOR = 3.50, 95% CI: 2.10–5.85), good system quality (AOR = 2.28, 95% CI: 1.38–3.78), adequate computer access (AOR = 2.06, 95% CI: 1.23–3.45), good system usability (AOR = 1.96, 95% CI: 1.20–3.19), and high eHealth literacy (AOR = 1.74, 95% CI: 1.05–2.89).

**Conclusion:** Satisfaction with LHIMS was moderate, influenced by both system-related and user-related factors. Strengthening information and system quality, enhancing usability, expanding computer access, and improving eHealth literacy are essential to optimising provider satisfaction and sustaining EMR use in Ghana.

## Introduction

Electronic Medical Record (EMR) systems are computerised platforms designed to collect, store, and manage patient health information (1). The system typically captures sociodemographic data, insurance details, medication history, allergies, laboratory results, immunisation records, and hospitalisation history, all safeguarded by strict security measures to ensure confidentiality (2). The concept of digital health records emerged in the 1960s, but large-scale adoption gained momentum in the 1990s, driven by advances in information technology and health informatics(3). In the 21^st^ century, particularly over the last decade, EMRs are recognised globally as essential for health sector reforms, with more than 90% of hospitals in high-income countries, such as the United States, using certified systems (4). Significant progress has also been made in enhancing adoption in low-and middle-income countries. At the policy level, the WHO Global Strategy on Digital Health 2020–2025 and the Sustainable Development Goals (SDG 3.8 on universal health coverage) position EMRs as critical for modernising health systems and improving population health(5,6)

EMR systems provide wide-ranging benefits. Clinically, they improve adherence to treatment guidelines, enhance preventive care, reduce medication errors, and ensure accurate and legible documentation(1,5). Operationally, the system streamlines workflows by eliminating redundant paper-based processes, supports real-time access to patient information, and enhances communication among multidisciplinary teams (7). At the organisational level, EMRs improve efficiency, reduce costs, and contribute to the sustainability of healthcare institutions, thereby enhancing patient trust and service uptake (8,9). In recognition of these benefits, international actors such as the World Health Organisation and the World Bank have actively promoted EMR adoption, particularly in sub-Saharan Africa, to improve health service delivery and strengthen health system performance (6,10).

Despite their promise, EMR implementation remains constrained in resource-limited settings. Financial and technical challenges, such as high system costs, inadequate infrastructure, and shortages of skilled personnel, pose significant barriers(11,12). Organisational bottlenecks, including weak policy frameworks, poor coordination, and inadequate technical maintenance, undermine sustainability(13). Furthermore, user-related factors—such as limited digital literacy, poor system usability, insufficient training, and a lack of end-user involvement in system design—often reduce provider confidence and hinder long-term use(7). The challenges underscore the pivotal role of healthcare providers, who are the primary end-users, in ensuring the successful adoption and sustainability of EMR systems.

In Ghana, health facilities historically relied on paper-based records until electronic medical record (EMR) systems were introduced to improve efficiency, accuracy, and data management(14,15). Early initiatives, such as the Hospital Administration Management System (HAMS) and JMeD, as well as ZenMedic, were introduced. However, these systems were privatised, expensive, fragmented, and lacked interoperability(16). To unify and scale the approach, the Ministry of Health (MoH) launched the National eHealth Strategy (2010–2017), which provided a roadmap for digitising health information systems, improving interoperability, and transitioning away from paper records (15). Building on this, the Digital Health Roadmap (2017–2024) identified the Lightwave Health Information Management System (LHIMS) as the core platform to strengthen service delivery and health system efficiency (14).

The operationalisation of the LHIMS began in 2017, with a pilot implementation in 2018 and its subsequent national approval in 2019 (14). Since then, LHIMS has expanded rapidly, now covering about 61% of public healthcare facilities and capturing the health records of nearly 21 million Ghanaians. The system is operational in all teaching and regional hospitals, with coverage also extending to many district hospitals, polyclinics, and health centres nationwide (17,18). Despite these achievements, challenges persist. Unreliable electricity, weak internet connectivity, inadequate technical support, and inconsistent user training continue to undermine system performance and reduce provider confidence(19,20). There have also been concerns about the lack of involvement and input from healthcare providers in the design and development of the system. Hence, key clinical functions are not appropriately captured in the system (21). These barriers directly influence both acceptability and sustainability of the system.

Addressing the challenges to utilization and sustainability of EMRs requires a clear understanding of healthcare providers’ satisfaction with the implemented EMRs and the determinants of that satisfaction. Providers’ satisfaction influences not only their willingness to adopt and use the system but also the long-term sustainability of digital health initiatives. However, research in Ghana has largely focused on other themes such as knowledge (22), utilisation (23,24), implementation (25), and post-implementation challenges(19) rather than satisfaction. Where satisfaction has been studied, it has predominantly concerned patients(26) or health leaders (21). Empirical research focusing on healthcare providers remains limited to the Central Region (27). No evidence exists from the Ashanti Region, despite it having the largest number of healthcare facilities in Ghana and one of the most extensive implementations areas of the LHIMS(14). This absence of provider-focused research in the Ashanti Region creates a significant evidence and contextual gap. Since healthcare providers’ satisfaction is a decisive factor in determining the acceptance, practical use, and sustainability of EMR systems, it is vital to identify both their satisfaction levels and the factors influencing them. To address this gap, the present study evaluated healthcare professionals’ satisfaction with the EMR system and analysed the determinants of their satisfaction in the Ashanti Region of Ghana.

## Materials and Methods

### Study Design and Period

A multicenter, analytical, cross-sectional study was conducted from 28^th^ August to 30^th^ October 2024. This design was appropriate, as it enabled the assessment of healthcare providers’ satisfaction with the LHIMS, their perceptions of its qualities, and the determinants of satisfaction at a single point in time. This design is consistent with approaches widely applied in health information research to establish statistical associations between system features and user outcomes, while minimising the influence of longitudinal variation (5,10,13).

### Study Area

The study was conducted in public healthcare facilities implementing LHIMS in the Ashanti Region of Ghana. The Ashanti Region is one of Ghana’s sixteen administrative regions, with Kumasi as its capital. The region lies in the middle belt of the country, specifically between longitudes 0.15^°^W and 2.25°W and latitudes 5.50°N and 7.46°N. The region encompasses a total land surface area of 24,389 square kilometres, accounting for 10.2% of Ghana’s total land area(28)

The Ashanti region is one of Ghana’s most populous and socioeconomically vibrant regions. It serves as a central referral hub for the middle and northern belts of the country. Healthcare in the region is organised into a tiered system comprising a teaching hospital (1), a regional hospital (1), district hospitals (26), polyclinics (8), health centres (162), and community-based health planning and services (CHPS) compounds (181) and several private healthcare facilities. With over 1,800 health facilities, it is one of the most densely served areas in the country and plays a central role in national health service delivery(29).

As part of Ghana’s eHealth strategy, the Ashanti Region has played a central role in the progressive rollout of the Lightwave Health Information Management System (LHIMS), the national electronic medical records platform. Implementation followed a phased approach, with teaching and regional facilities being the first to adopt LHIMS (2019–2020), followed by district hospitals (2021–2022), and then primary-level facilities, such as polyclinics and health centres (2022–2023) (14). This staged implementation created natural variation in LHIMS maturity across levels of care, making the region an ideal setting for assessing user satisfaction.

Three facilities representing different levels of healthcare delivery were selected for this study. Kumasi South Regional Hospital (KSRH), located in the Asokwa Municipality of Kumasi, serves as the designated regional hospital for the Ashanti Region. It provides specialised and referral services, supports medical and nursing training, and has a bed capacity of about 120, with departments covering internal medicine, surgery, paediatrics, obstetrics and gynaecology, and accident and emergency. It was one of the first regional hospitals to implement LHIMS, starting around 2019 (14).

Suntreso Government Hospital (SGH), situated in the Kumasi Metropolis, functions as a district hospital, providing outpatient and inpatient services that encompass general medicine, maternal and child health, surgery, and laboratory support. With a bed capacity of about 60, SGH plays a vital role in delivering secondary-level care to surrounding urban communities. LHIMS was introduced at SGH between 2021 and 2022, in line with the phased rollout to district-level facilities(14).

Asuofua Polyclinic, situated in the Atwima Nwabiagya District, is a primary-level facility that offers outpatient care, maternal and child health services, preventive care, and limited inpatient care. The polyclinic serves as the first point of contact for healthcare in its catchment area.

LHIMS was implemented at Asuofua Polyclinic in 2022–2023, during the expansion to primary-level facilities, thereby linking grassroots care into Ghana’s national digital health infrastructure.

By selecting facilities from three levels of healthcare delivery, tertiary/regional, district, and primary, the study ensured representativeness and captured diverse provider experiences with LHIMS across different institutional contexts. This design also enabled meaningful comparisons of satisfaction across facilities with varying mandates, capacities, and durations of exposure to the system.

### Study Population

The source population comprised all healthcare professionals working in public health facilities in the Ashanti Region of Ghana. The study population consisted of healthcare providers employed in these facilities who actively used the Lightwave Health Information Management System (LHIMS). Healthcare providers, defined as clinical staff directly engaged in patient care, are the frontline users of LHIMS in documentation, prescribing, dispensing, and managing laboratory results. Their satisfaction is central to the system’s effective use and sustainability.

### Inclusion and Exclusion Criteria

Inclusion was restricted to providers who had password access and actively used LHIMS, ensuring authorised access and direct experience with the system.

Exclusion applied to healthcare providers who were on leave (annual, maternity, or other forms) during the study period, as confirmed from the human resource leave roster, and those with less than three months of LHIMS experience, to minimise bias from limited exposure and ensure adequate familiarity with system functions.

### Sample Size Determination

The sample size was determined using Cochran’s formula for estimating a single population proportion (30) assuming a 95% confidence interval, 5% significance level (𝛼) *(two-sided),* 5% margin of error, and an expected user satisfaction with electronic medical records of 84.8%(13). To account for variability introduced by the multistage sampling design, a design effect (D_eff) of 2.0 was applied. A 10% adjustment was added to compensate for potential non-response. The final estimated sample size was 436.

The sample size (n) was calculated as follows;

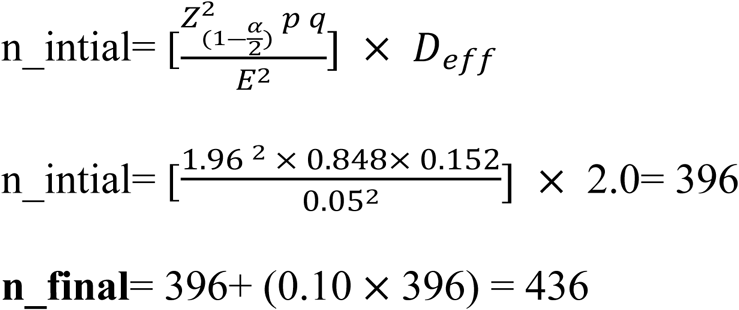

where n_intial is the estimated sample size, p is the proportion of user satisfaction, Z **_(1-_**𝜶/2) is the Z-statistic value at the 95% confidence level and 5% significance level (α = 0.05) for 2 tails, which is 1.96, E is the margin of error 5% (0.05), and 𝐷_𝑒𝑓𝑓_ is the design effect of 2.0 and n_final is final sample size.

### Sampling Technique

A multistage stratified sampling technique was employed to ensure fair representation of healthcare providers using LHIMS across different levels of care. The process involved two stages: facility selection and participant selection.

### Selection of Facilities

All healthcare facilities in the Ashanti Region with functional LHIMS were stratified into three levels of care: tertiary/regional, district, and primary. From each stratum, one facility was randomly selected using the RANDBETWEEN function of Microsoft Excel version 2020 to reflect both the level of care and the stage of LHIMS adoption. Accordingly, Kumasi South Regional Hospital (regional), Suntreso Government Hospital (district), and Asuofua Polyclinic (primary) were chosen for the study.

### Selection of Participants

The Health Information Departments of the selected facilities provided rosters of healthcare providers with access to LHIMS. The rosters initially included 672 providers at Kumasi South Regional Hospital, 523 at Suntreso Government Hospital, and 232 at Asuofua Polyclinic. After applying the study’s eligibility criteria, the sampling frame was reduced to 645, 499, and 219 providers, respectively.

The total sample size of 436 participants was proportionally allocated across the three facilities based on the size of their eligible populations: 206 from Kumasi South Regional Hospital, 160 from Suntreso Government Hospital, and 70 from Asuofua Polyclinic.

Within each facility, simple random sampling was then applied. Unique identification numbers were assigned to all eligible providers, and random numbers were generated using the RAND function in Microsoft Excel to select participants. A reserve list (10–15% of the required sample per facility) was also prepared to replace non-responders after two unsuccessful contact attempts.

Finally, selected providers were approached in their offices or wards, and eligibility was reconfirmed; the objectives of the study were then explained. Written informed consent was obtained before questionnaire administration. A flow diagram of the sampling process is presented in Fig 1.

**Fig 1:**
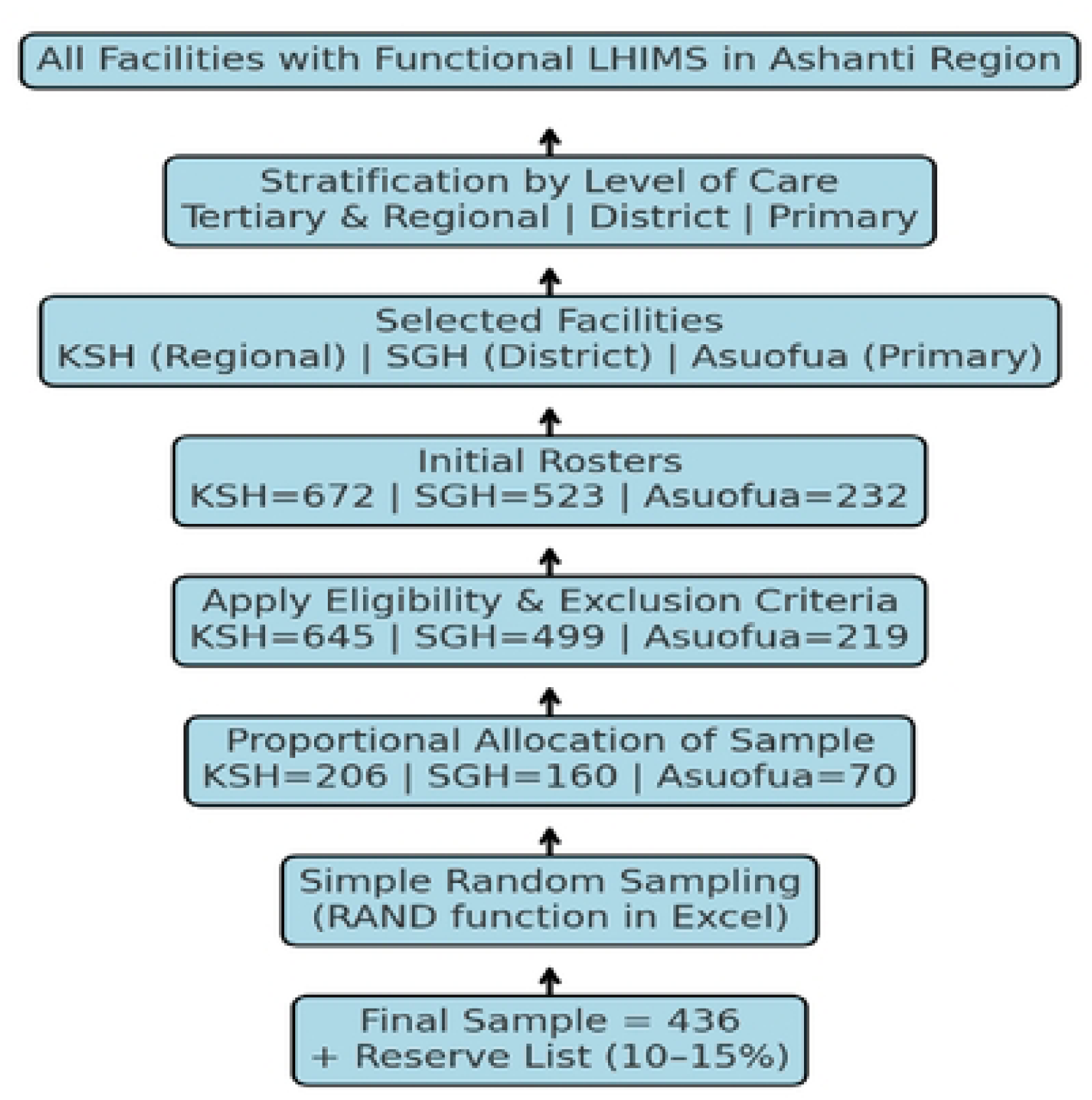
Flow diagram of the sampling process

### Instrumentation

A structured, self-administered questionnaire was used to collect data from the healthcare providers. The instrument was developed from standardised and previously validated tools, such as the eHealth Literacy Scale (eHEALS) (31) and the System Usability Scale (SUS) (32). These were supplemented with items adapted from the literature (5,10,33) as well as researcher-designed socio-demographic questions. The questionnaire was prepared in English and organised into the following sections: socio-demographic characteristics, user background, user satisfaction, and perceived technological qualities.

### Study Variable and Measurement Dependent Variable

#### User Satisfaction

measured healthcare providers’ overall contentment with the EMR system. It was assessed with seven items adapted from Dubale et al. (5), based on the *user satisfaction* construct of the DeLone & McLean IS Success Model. Each item was scored on a 5-point Likert scale, ranging from 1 (strongly disagree) to 5 (strongly agree). Composite scores ranged from 7 to 35 points, with higher values representing greater satisfaction. Normality was assessed using the Shapiro–Wilk test. With p < 0.05, data were considered non-normally distributed, and scores were dichotomised at the median split. Participants with scores equal to or above the median were classified as satisfied. Dubale et al. (5) reported a Cronbach’s alpha value of 0.78.

### Independent Variables

#### Socio-demographic Characteristics

This section used seven researcher-designed items to assess age, gender, professional category (nurse, physician, physician assistant, midwife, or laboratory scientist), years of professional experience, and healthcare facility.

#### User background

This section comprised four constructs: professional training, access to computers, eHealth literacy, and computer literacy.

- **Professional training** was measured with one (1) dichotomous item (yes/no) asking whether the healthcare provider had received training on electronic medical records from the Health Information Department.
- **Access to computers** was evaluated with one (1) item rated as adequate, moderate, or inadequate.
- **eHealth literacy** was assessed using the 8-item eHealth Literacy Scale (eHEALS)(31).

Each item was rated on a 5-point Likert scale (1 = strongly disagree to 5 = strongly agree), resulting in scores from 8 to 40. Higher scores indicated greater literacy. Normality was tested with the Shapiro–Wilk test, and since the data were non-normally distributed (p < 0.05), the median split was applied. Participants who scored equal to or above the median were categorised as having high eHealth literacy. The original eHEALS reported a Cronbach’s alpha of 0.88 (31).

- **Computer literacy** was assessed using four items adapted from Dubale et al. (5) rated on a 5-point Likert scale (1 = strongly disagree to 5 = strongly agree). Composite scores ranged from 4 to 20, with higher scores reflecting better literacy. The median split was applied to classify participants into two groups: those with high computer literacy and those with low computer literacy.

### Perceptions of the Technological Qualities

#### System Quality

was assessed based on the perceived reliability, response time, and overall functionality of the EMR system. It was measured using six items adapted from Dubale et al. (5) rated on a 5-point Likert scale (1 = strongly disagree to 5 = strongly agree). Composite scores ranged from 6 to 30, with higher scores indicating better system quality. The Shapiro–Wilk test confirmed non-normal distribution, and participants were categorised into good versus poor system quality groups using the median split. Scores at or above the median were considered good perceived system quality.

#### Information Quality

referred to the accuracy, completeness, timeliness, and relevance of EMR data. It was measured using seven items adapted from the DeLone & McLean IS Success Model and prior studies (5,10). Each item was rated on a 5-point Likert scale, resulting in scores ranging from 7 to 35. Higher scores reflected better information quality. The Shapiro–Wilk test indicated non-normality, and participants were classified into two groups based on good and poor information quality using a median split.

#### Service Quality

was assessed based on the responsiveness and dependability of technical and infrastructural support for the EMR system. It was measured using nine items adapted from the D&M framework and Dubale et al. (5). The items were rated on a 5-point Likert scale, with a composite score ranging from 9 to 45. Higher scores indicated stronger service quality. Following the Shapiro–Wilk test, the median split was applied to categorise participants into good and poor service quality groups. Scores at or above the median were considered “good” service quality.

#### System Usability

was assessed using the validated System Usability Scale (SUS) (32). The SUS includes ten items measuring ease of navigation, user-friendliness, and efficiency of the EMR interface. Questions ***1, 3, 5, 7, and 9*** were positive statements, while items ***2, 4, 6, 8, and 10*** were negative statements. Each item was rated on a 5-point Likert scale, yielding scores ranging from 10 to 50. Higher scores indicated greater usability. Based on the Shapiro–Wilk test, data were non-normally distributed, and participants were classified into good or poor usability groups using the median split. Scores at or above the median were considered good system usability. The original SUS has a reliability coefficient of 0.90 (32)

#### Data Collection Procedures

Data were collected by six trained research assistants with backgrounds in nursing and physician assistantship, under the supervision of a medical officer, an MPhil nursing student, and the principal investigator *(two assistants and one supervisor at each facility)*.

Eligible healthcare providers identified through the sampling procedure were contacted individually in their respective offices or wards. At the initial contact, the purpose of the study, its objectives, and procedures were explained, and written informed consent was obtained before enrollment. To maintain anonymity during fieldwork, each participant was assigned a unique study identification number, which replaced names on all instruments.

Recruitment outcomes were documented in a logbook. Providers who declined participation were recorded, while those who could not be reached after two separate attempts were classified as non-responders. Replacements were drawn sequentially from a pre-generated reserve list within the same facility. To minimise disruption to clinical duties, data collection was scheduled during less busy hours, and participants completed the questionnaire at a convenient time within their work units. Questionnaires were self-administered, with research assistants available to provide clarification when needed. Completed forms were checked on-site for accuracy and completeness, with immediate follow-up when omissions were found.

Supervisors provided on-site oversight across facilities and reviewed completed questionnaires on a daily basis. The research team held daily debriefing sessions to resolve challenges and harmonise procedures. At the end of each week, questionnaires were securely transferred to the principal investigator for storage and preparation for data entry.

#### Quality Assurance: Validity and Reliability

The quality of the study instrument was assured through validity and reliability checks. Content validity was achieved by adopting standardised and widely used measures, including the eHealth Literacy Scale (eHEALS) and the System Usability Scale (SUS), supplemented with validated items adapted from the literature (5,10,31,32).

Face validity was confirmed by the Trainer of Trainees of the Lightwave e-Health Solutions, who reviewed the questionnaire for clarity, comprehensiveness, and contextual relevance.

A pretest was conducted at Effiduase Government Hospital to confirm applicability. Based on participant feedback, one item related to District Health Information Management System (DHIMS) training was removed because it was limited to middle-level health managers and not associated with the utilisation of LHIMS (34).

Construct validity was further examined using exploratory factor analysis (EFA). Prior to extraction, the Kaiser–Meyer–Olkin (KMO) measure of sampling adequacy and Bartlett’s test of sphericity were applied to confirm suitability for factor analysis. The KMO value was high (0.889), indicating that the data were suitable for factor analysis. Bartlett’s test of sphericity was statistically significant (p < 0.001), confirming that the correlation matrix was appropriate for factor extraction (35). The EFA revealed that all constructs had eigenvalues greater than 1, and extracted factors explained a cumulative variance of 61.3%, exceeding the 50% minimum benchmark recommended in health sciences. Item loadings ranged from 0.39 to 0.64. The lowest loading (0.39) was slightly below the conventional cutoff of 0.40 but was retained due to its theoretical relevance and overall contribution to the construct. Most items exceeded the 0.40 threshold, indicating acceptable construct validity (See Table 1.)

**Table 1:** Validity and Reliability of Measurement Tools.

| statistic | Value |
| --- | --- |
| Kaiser-Meyer-Olkin (KMO) | 0.889 |
| Bartlett's Test of Sphericity (p value) | <0.001 |
| Number of Extracted components<br>(Eigenvalue>1) | All constructs (multi-items) |
| Cumulative variance explained | 61.3% |
| Range of Factor Loadings | 0.385-0.640 |
| Cronbach alpha | User satisfaction=0.811, service quality=0.869,<br>information quality=0.862, system quality=0.747,<br>system usability=0.740. eHealth literacy=0.651,<br>computer literacy=0.641 |

Reliability was assessed using Cronbach’s alpha to estimate internal consistency. Cronbach’s alpha values ranged from 0.64 to 0.87, indicating acceptable internal consistency for exploratory research (36) (See Table 1.)

To enhance procedural reliability, all research assistants and supervisors underwent a two-day training session covering study objectives, ethical conduct, and standardised administration techniques before data collection commenced. Overall, the instruments were both valid and reliable

#### Ethical Considerations

Ethical approval for the study was granted by the Committee on Human Research, Publications, and Ethics (CHRPE) at the Kwame Nkrumah University of Science and Technology **(reference number: [CHRPE/AP/977/24])**. Administrative approval was obtained from the management of the selected hospitals before data collection.

Participation was entirely voluntary. Written informed consent was obtained from all participants after they were informed of the study objectives, procedures, potential risks, and benefits. Participants were informed of their right to decline or withdraw at any point without consequences.

To ensure confidentiality and anonymity, no personal identifiers, such as names, were recorded. Unique study identification numbers **(Questionnaire ID**) were used on all instruments. Completed questionnaires were stored securely in locked cabinets and later transferred to password-protected electronic databases accessible only to the research team.

The study posed minimal risk, as it involved no invasive procedures. The potential benefit lies in generating evidence that could inform improvements in the implementation of electronic medical records and healthcare delivery.

## Data Analysis

Data were entered and cleaned in Microsoft Excel 2020 for Mac and exported to IBM SPSS Statistics version 25 for analysis. Descriptive statistics were used to summarise sociodemographic characteristics, user background, perceptions and satisfaction. Categorical variables were presented as frequencies and percentages, while continuous variables were summarised using means with standard deviations (SD) if normally distributed, or medians with interquartile ranges (IQR) if skewed.

Logistic regression analysis was applied to identify determinants of user satisfaction with the electronic medical records system. Bivariate logistic regression was first performed to estimate crude odds ratios (COR) with 95% confidence intervals (CI). Variables with a p-value < 0.25 in the bivariate analysis were considered for entry into the multivariable model, as this liberal cutoff helps avoid prematurely excluding potential predictors that may become significant after controlling for confounders (37).

Multivariable analysis was conducted using hierarchical logistic regression. Two models were specified. **Model 1** included theory-driven constructs from the DeLone and McLean Information Systems Success Model, namely system quality, information quality, service quality, training, computer literacy, and eHealth literacy, regardless of their bivariate significance. **Model 2**, the final explanatory model, extended Model 1 by incorporating contextual variables that showed at least a marginal association in the bivariate analysis (p < 0.25), including sociodemographic and professional characteristics (age, sex, cadre, and years of practice, etc.).

Model diagnostics were performed during the multivariable stage. Multicollinearity was assessed using variance inflation factors (VIF), with a cut-off of 5. “Facility level” and “computer access” were found to be highly correlated. The facility level was excluded, while computer access was retained due to its stronger theoretical and practical relevance to information system success(54). Model fit was adequate, as indicated by the Hosmer–Lemeshow goodness-of-fit test (p = 0.537). The discriminative ability of the final model was good, with an area under the receiver operating characteristic curve (AUROC) of 0.784. These diagnostics confirmed that the final explanatory model was statistically robust and provided a reliable basis for identifying independent determinants of user satisfaction.

Determinants were defined as variables that remained statistically significant in the final multivariate model at p < 0.05 and were presented as adjusted odds ratios (AORs) with 95% confidence intervals (95%)

## RESULTS

### Sociodemographic and Professional Characteristics of the Healthcare Providers (N=374)

Table 2 presents the socio-demographic and professional characteristics of the participants. Out of the 438 participants surveyed, 374 responses were valid for data analysis, resulting in a response rate of 88.4%. The mean age was 33 years (*SD* = 5.70), with 114 (38.4%) aged above 35 years. Two-thirds of the participants (n = 247, 66%) were female, while more than half (n = 220, 58.8%) were married. The majority of participants (n = 307, 82%) were Christians. Almost half of the participants (n = 183, 49%) were recruited from Kumasi South Regional Hospital, and nurses represented the largest professional group (n = 174, 49.5%).

**Table 2:** Sociodemographic and Professional Characteristics of the Healthcare Providers (N=374)

| Variable | Category | Frequency (n) | Percentage (%) |
| --- | --- | --- | --- |
| **Age (years)** |  |  |  |
| *Mean age =33 years*<br>*(SD=5.70)* | 20-29 | 111 | 29.6 |
|  | 30-35 | 119 | 32.0 |
|  | Above 35 years | 144 | 38.4 |
| **Gender** |  |  |  |
|  | Male | 127 | 34.0 |
|  | Female | 247 | 66.0 |
| **Religion** |  |  |  |
|  | Christian | 307 | 82.0 |
|  | Muslim | 67 | 18.0 |

| Marital status |  |  |  |
| --- | --- | --- | --- |
|  | Single | 145 | 38.8 |
|  | Married | 220 | 58.8 |
|  | Others (divorced, widowed) | 9 | 2.40 |
| Professional category |  |  |  |
|  | Nurse | 174 | 46.5 |
|  | Midwives | 94 | 25.1 |
|  | Doctor | 54 | 14.4 |
|  | Pharmacist | 35 | 9.5 |
|  | Other (PA, medilab, etc.) | 17 | 4.5 |
| Years of Practice |  |  |  |
| *Mean= 6.33 (SD=4.07)* | 1-5 | 195 | 52.1 |
|  | 6-10 | 120 | 32.1 |
|  | Above 10 | 59 | 15.8 |
| Health facility |  |  |  |
|  | KSRH | 183 | 49.0 |
|  | SGH | 128 | 34.0 |
|  | Asuofua Polyclinic | 63 | 17.0 |

*KSRH=Kumasi South Regional Hospital, SGH=Suntreso Government Hospital,*
*PA=Physician Assistant, Medilab=Medical Laboratory Scientist*

### User background: Professional Training, Computer Literacy, and Proficiency

Table 3 presents the results regarding user background in terms of professional training, computer literacy, and proficiency. The majority of participants (n = 351, 94.0%) had received training on LHIMS, while exactly half (n = 188, 50%) reported having fair access to computers at their facility. Additionally, 262 (70%) demonstrated high computer literacy, whereas 199 (53.2%) had low eHealth literacy.

**Table 3:** User background: Professional Training, Computer Literacy, and Proficiency (N=374)

| Variable | Category | Frequency<br>(n) | Percentage<br>(%) |
| --- | --- | --- | --- |
| Access to computers in your hospital | Adequate | 164 | 44.0 |
|  | Fair | 188 | 50.0 |
|  | Inadequate | 22 | 6.0 |
| LHIMS Training | Yes | 351 | 94.0 |
|  | No | 23 | 6.0 |
| Computer literacy | High | 262 | 70.0 |
|  | Low | 112 | 30.0 |
| eHealth Literacy | High | 175 | 46.8 |
|  | Low | 199 | 53.2 |

### Healthcare Providers’ Satisfaction with Light Wave Health Information Management System (LHIMS)

The overall median satisfaction score was 29 (*IQR*: 27–32). Based on the median cut-off, 213 healthcare providers (57%, 95% CI: 52%–62%) were categorised as satisfied with the LHIMS (<u>see</u> Fig 2).

**Fig 2:**
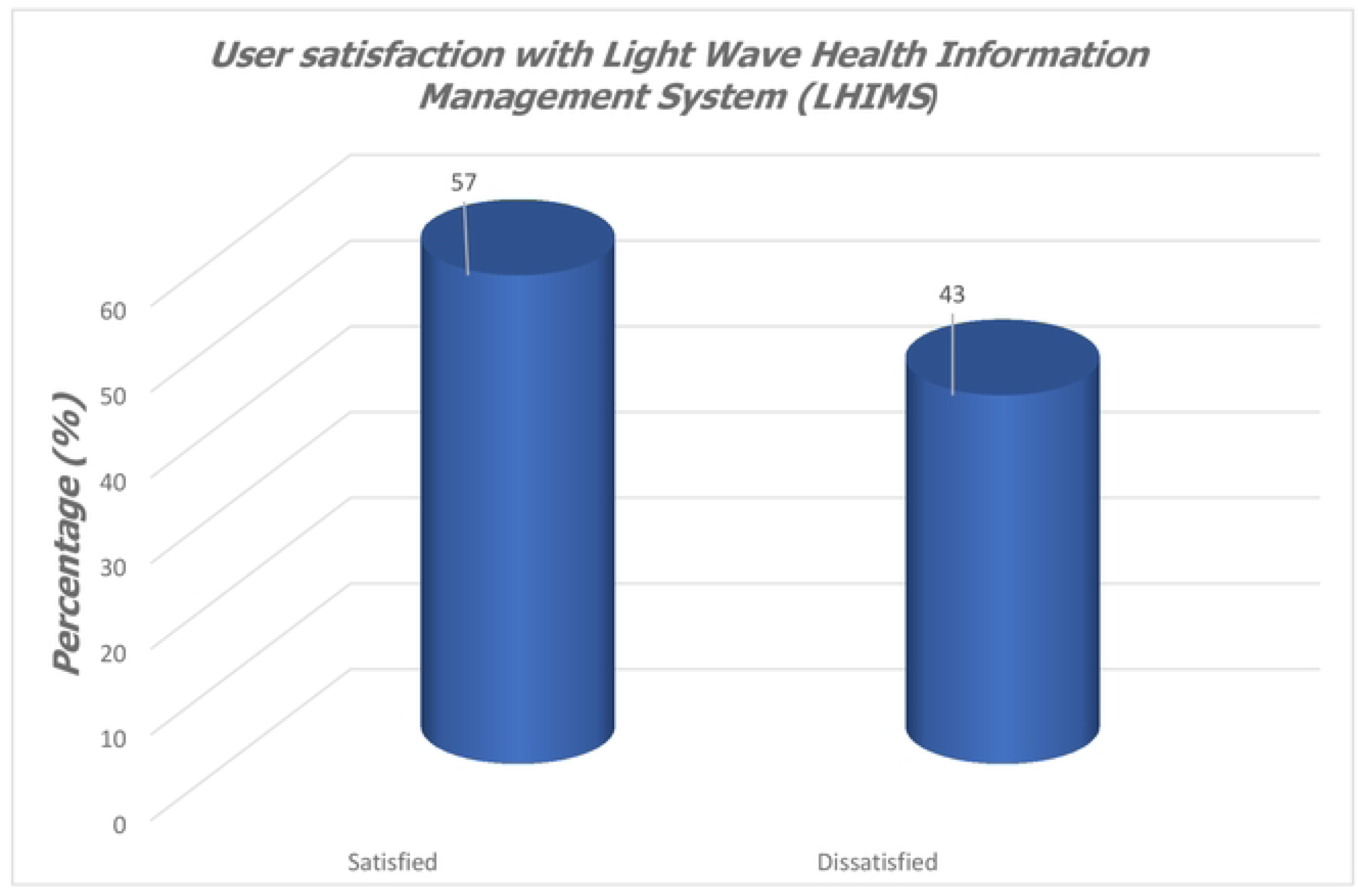
Overall Healthcare Providers’ Satisfaction with Light Wave Health Information Management System (LHIMS) (N=374, Median=29 (IQR=27–32)

At the item level, the highest-rated domain was the preference for the LHIMS over the paper-based records system (mean = 4.35, SD = 0.77), followed by the perception that the LHIMS helps healthcare providers complete their work faster (mean = 4.27, *SD* = 0.61). Satisfaction with the system’s contribution to improved information quality was also high (mean = 4.22, *SD* = 0.72). (<u>see</u> Table 4)

**Table 4:** User satisfaction with Light Wave Health Information Management System (LHIMS) (N=374)

| Variable | Mean | SD |
| --- | --- | --- |
| LHIMS help me to finish my work faster | 4.27 | 0.61 |
| My performance and productivity have improved due to LHIMS | 4.07 | 0.71 |
| The system has positive impact on quality of care | 4.19 | 0.65 |
| Patient's safety has improved due to LHIMS | 4.06 | 0.72 |
| Information quality has improved due to LHIMS | 4.22 | 0.72 |
| LHIMS worth the time and effort required to use it | 4.20 | 0.68 |
| Prefer the LHIMS system to the paper record | 4.35 | 0.77 |
| **Overall Users' Satisfaction** | **29.35** | **3.34** |

Although satisfaction was generally positive, the lowest rating was observed for the system’s impact on quality of care (mean = 4.19, SD = 0.65), performance and productivity (mean = 4.07, *SD* = 0.61), and patient safety (mean = 4.06, *SD* = 0.72) (see Table 4).

Detailed results regarding healthcare providers’ satisfaction with LHIMS are presented in Table 4 and Fig 2.

### Perceived Qualities of the Light Wave Health Information Management System (LHIMS)

Table 5 presents results regarding healthcare providers’ perceptions of the qualities of LHIMS.

**Table 5:** Perceived Qualities of the Light Wave Health Information Management System.

| Variable | Median<br>(IQR) | Category | Frequency<br>(n) | Percentage<br>(%) |
| --- | --- | --- | --- | --- |
| System Usability | 32 (30-36) | Good | 215 | 57.5 |
|  |  | Poor | 159 | 42.5 |
| System quality | 25 (23-27) | Good | 188 | 50.3 |
|  |  | Poor | 186 | 49.7 |
| Service Quality | 34 (30-37) | Good | 196 | 52.4 |
|  |  | Poor | 178 | 47.6 |
| Information Quality | 29 (27-31) | Good | 194 | 52.0 |
|  |  | Poor | 180 | 48.0 |

More than half of the participants (n=215, 57.5%) considered the system’s usability to be good, and 188 (50.3%) rated the system’s quality as good. Similarly, 194 (52.0%) indicated good service quality, and 194 (52.0%) acknowledged good information quality.

### Bivariate and Multivariable Analysis of the Determinants of Healthcare Providers’ Satisfaction with LHIMS

In the unadjusted/bivariate analysis, **access to computers**, **computer literacy**, **eHealth literacy**, **perceived information quality**, **perceived system quality, perceived service quality**, and **perceived system usability** were all significantly associated with satisfaction at a significance level of P < 0.05 (see Table 6).

**Table 6:**
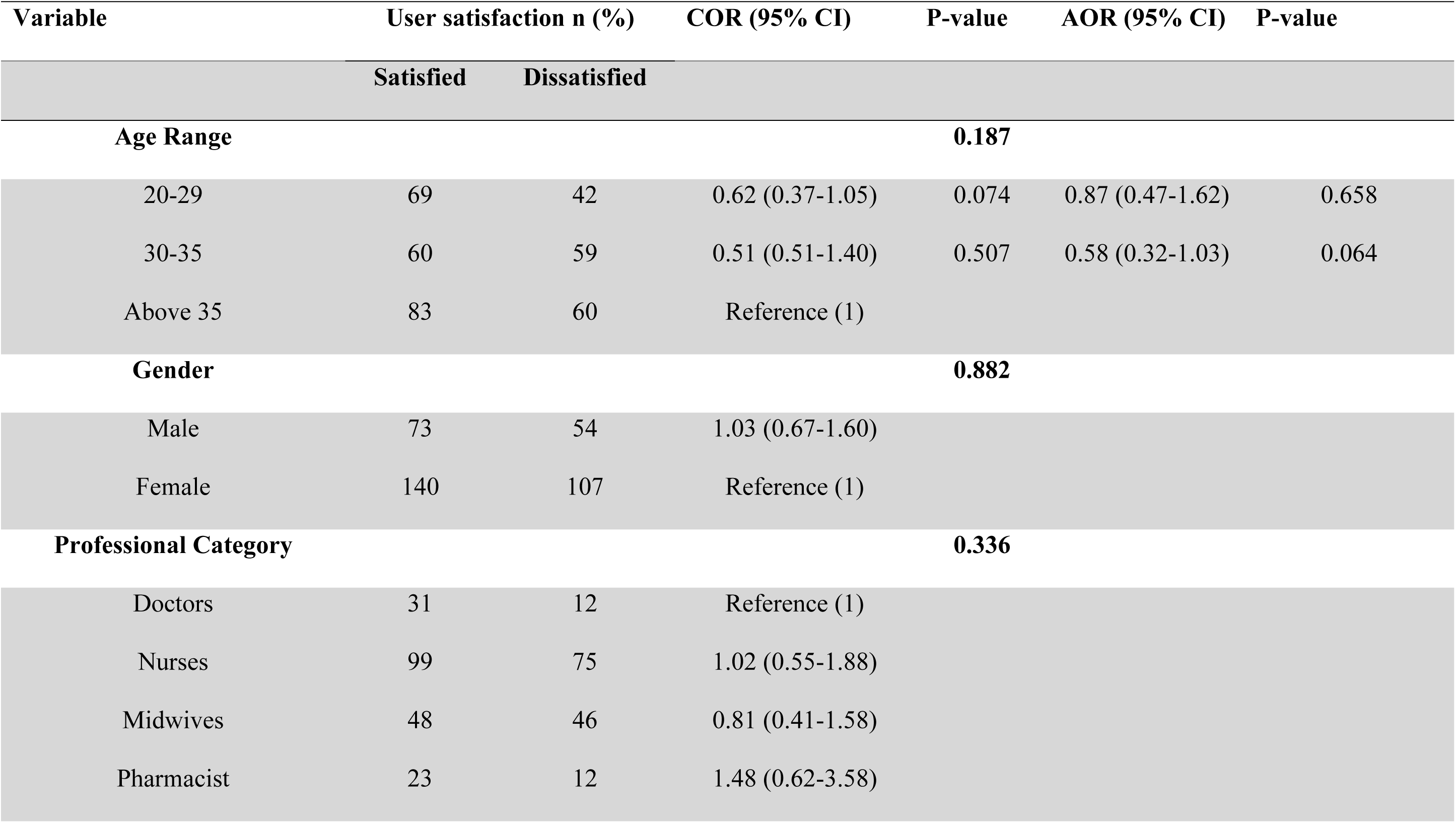
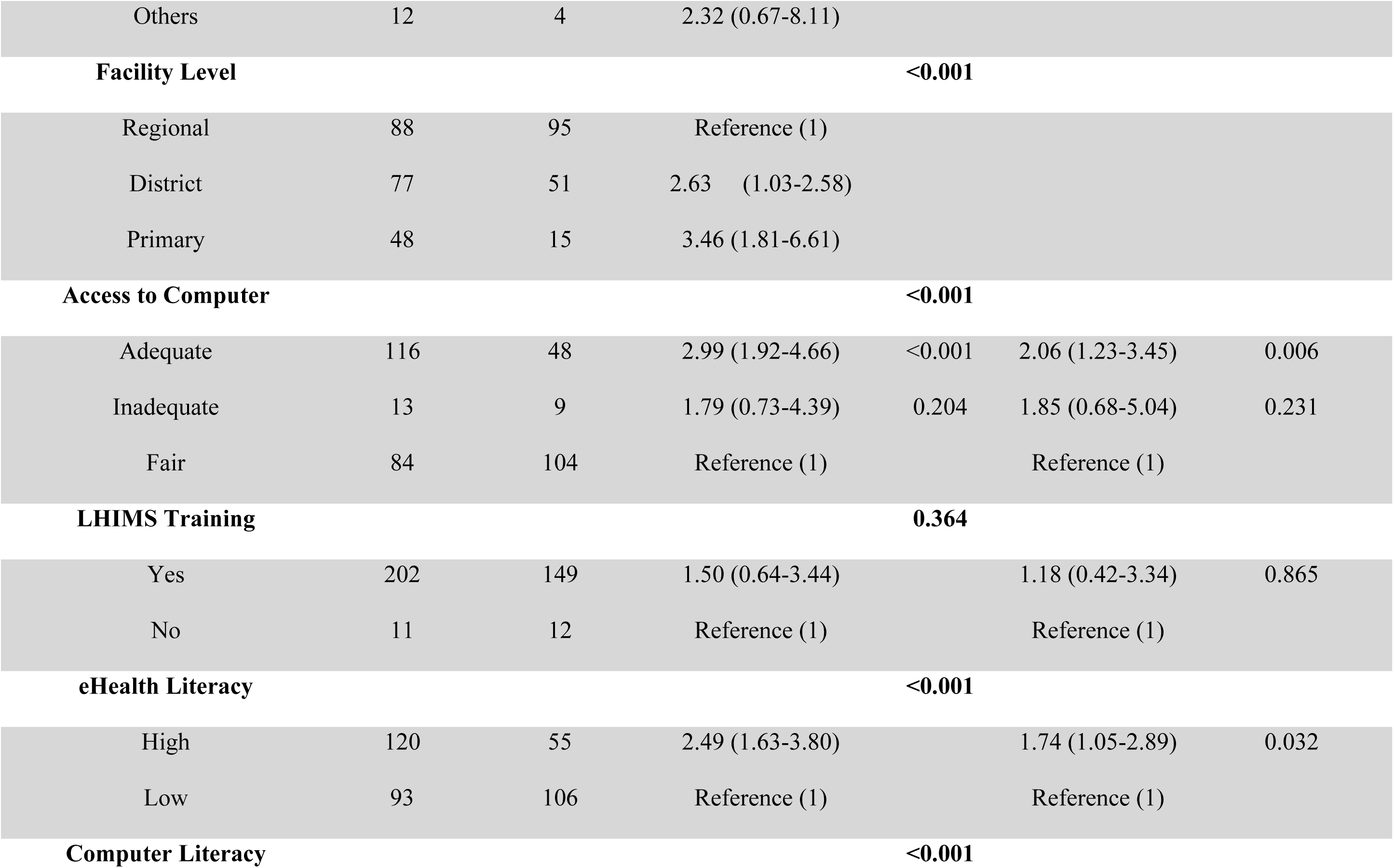
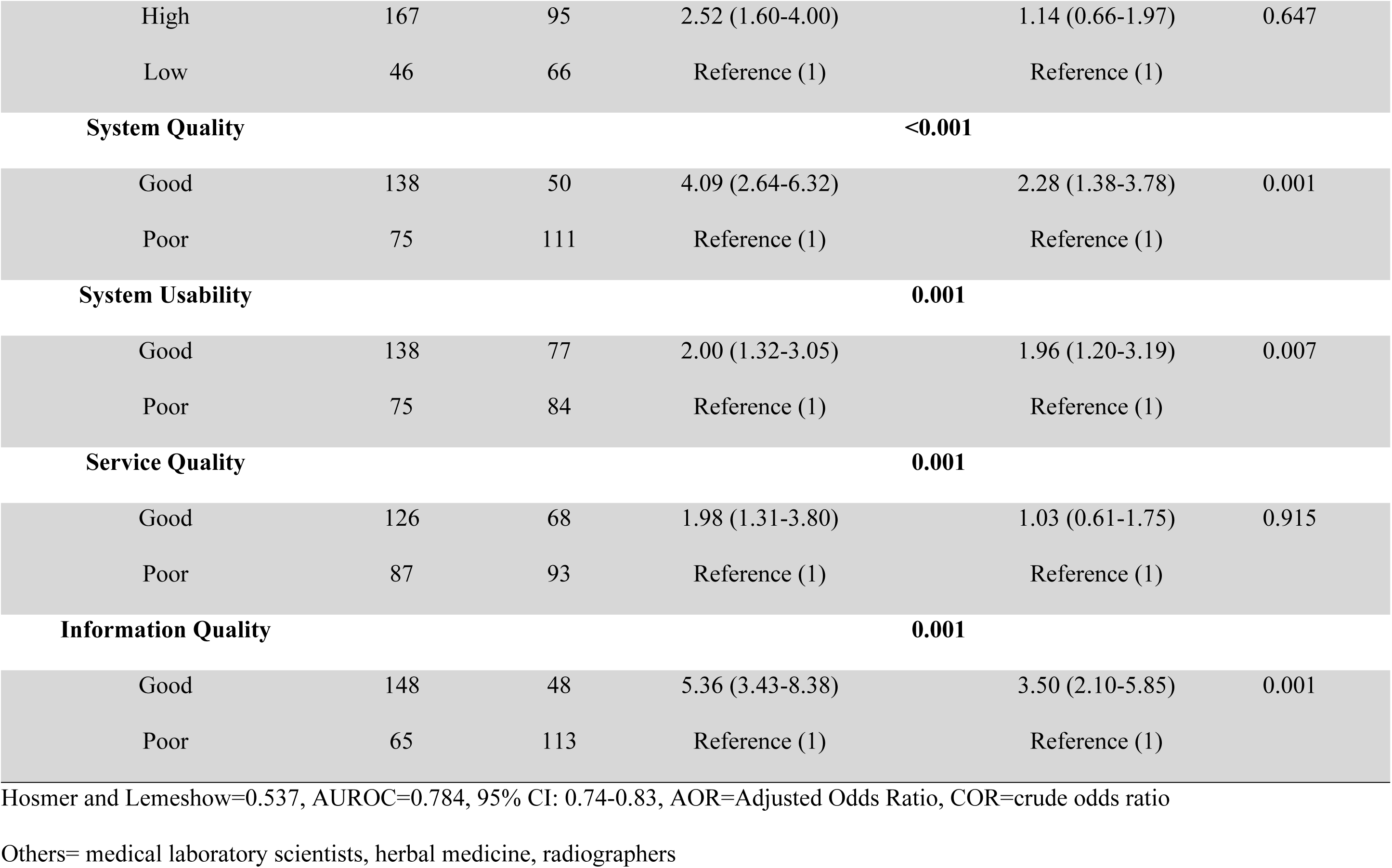
Bivariate and multivariable logistic regression analysis of the determinants of user satisfaction with the LHIMS.

In the adjusted or multivariable analysis, **perceived information quality**, **perceived system quality**, perceived **system usability**, **eHealth literacy**, and **computer access** remained significantly associated with healthcare providers’ satisfaction with the LHIMS after adjusting for possible confounders. Providers with **good perceived information quality** were over three times more likely to be satisfied with LHIMS (AOR = 3.50, 95% CI: 2.10–5.85). Good **perceived system quality** (AOR = 2.28, 95% CI: 1.38–3.78) and **adequate access** to computers (AOR = 2.06, 95% CI: 1.23–3.45) doubled the odds of satisfaction. Good perceived system usability (AOR = 1.96, 95% CI: 1.20–3.19) and high eHealth literacy (AOR = 1.74, 95% CI: 1.05–2.89) independently increased the odds of healthcare providers’ satisfaction. (<u>see</u> Table 6).

Although perceived service quality (AOR=1.98, 95% CI: 1.31-3.80) and computer literacy (AOR=2.52, 95% CI:1.60-4.00) were significant in the unadjusted analysis, these variables lost significance in the multivariable model (i.e., could not independently predict healthcare providers’ satisfaction after controlling for possible confounders). (<u>see</u> Table 6).

Moreover, LHIMS training and socio-demographic and professional factors, such as age, gender, professional category, and work experience, were not significantly associated with satisfaction. Detailed results are presented in Table 6

## DISCUSSION

### User Satisfaction with LHIMS

This study assessed healthcare providers’ satisfaction with the Lightwave Health Information Management System (LHIMS) in public facilities in the Ashanti Region. Overall satisfaction was moderate, with 57% of the participants reporting satisfaction. This suggests that while LHIMS has gained some acceptance among frontline providers, its full potential has not yet been realised. This finding is comparable to findings from Ethiopia, where 53% of providers in private health facilities expressed satisfaction with their electronic medical records system (5). The similarity of results may be attributed to methodological consistency, as both studies employed the same validated instrument, applied similar scoring approaches, and used median cut-offs to dichotomise satisfaction scores.

Contrastingly, studies from Saudi Arabia, the United States of America, and Canada have reported higher satisfaction levels (38–40). Structural and infrastructural disparities can explain these discrepancies. In high-resource settings, EMR systems are typically supported by multiple workstations per unit, stable internet connectivity, reliable electricity, and dedicated technical support. Such conditions enable providers to fully utilise EMR functionalities, including retrieving laboratory and radiological results, prescribing medications, and issuing treatment orders. In Ghana, however, infrastructural challenges, such as limited workstations, intermittent power supply, prolonged internet downtimes, and occasional service disruptions, undermine these processes and reduce user satisfaction (21).

Furthermore, Rahman et al. (13) and Maraga et al. (41) reported significantly higher satisfaction levels in Saudi Arabia (84%) and Rwanda (91%), respectively. The difference could be attributed to the system’s maturity. In their respective contexts, the EMR had been implemented for nearly a decade in Saudi Arabia and almost 15 years in Rwanda, allowing deficiencies to be corrected, repeated training to be provided, and user-driven upgrades to be introduced. In contrast, LHIMS has been operational for less than five years in the selected facilities, suggesting it is still in a mid-adoption stage. With further optimisation and sustained support, LHIMS could achieve higher levels of satisfaction and long-term sustainability.

Beyond the overall satisfaction, variations were observed across specific domains. Healthcare providers reported higher satisfaction with the quality of information and expressed a stronger preference for electronic records over paper-based documentation. These findings align with evidence from Norway, where EMR systems demonstrated superior information quality over handwritten records (42). The possible explanation could be that EMRs produce more precise, legible, and error-free documentation (43). System-generated outputs are structured and standardised, making them easier to interpret and share. These attributes enhance clinical decision-making and communication among healthcare providers, thereby explaining why they prefer LHIMS to paper-based documentation, which is often illegible and difficult to interpret.

Satisfaction with efficiency was also reported to be high, as providers indicated that LHIMS helped them complete their work faster. This finding is consistent with studies in Ethiopia and Saudi Arabia, where the use of EMRs improved efficiency by simplifying workflows and facilitating quicker access to patient data(6,10). A likely reason is that once information is entered into the system, it can be retrieved and reused instantly, unlike paper documentation that requires repeated entries and manual searching of patients’ folders.

Conversely, satisfaction was lower in domains related to patient safety and quality of care. One key feature of LHIMS is that it allows staff across units to navigate through patient information without restriction. While this design improves access, it also raises concerns about confidentiality, data security, and patient trust. Similar governance challenges have been highlighted in Ghana and other African contexts, where weak data protection frameworks and limited accountability mechanisms undermine user confidence in EMR systems(21,44).

Furthermore, consistent with other studies, field observation revealed users often spent extended periods working on the system, for documentation, retrieving laboratory results, pharmacy instruction etc., which reduces the time available for direct patient care (45). This shift in focus from the bedside to the computer station may have inadvertently contributed to lapses in care delivery and, in some cases, potential medical negligence. These factors therefore explain why satisfaction with LHIMs on quality of care and patients’ safety was relatively lower.

Overall, these findings suggest that while LHIMS improves documentation quality and efficiency, unresolved concerns about data security and bedside engagement continue to limit its overall impact on patient safety and quality of care.

### Determinants of Health Providers’ Satisfaction with LHIMS

The study analysed the determinants of healthcare providers’ satisfaction with LHIMS. User background factors were found to influence satisfaction. Access to computers emerged as a significant determinant, with providers who had adequate access reporting higher levels of satisfaction. This finding aligns with research from Ethiopia and Malaysia, where the adequate access to computers such as availability of more workstations and ownership of personal laptops and tablets, enhanced users satisfaction with the implemented EMR systems(10,46). This finding was expected because adequate computer access reduces competition for devices, ensures smooth workflow, and facilitates the timely completion of tasks such as data entry and retrieving laboratory results. Ensuring equitable distribution of computers across facilities is therefore vital to optimising the use and enhance users’ satisfaction with LHIMS in Ghana.

eHealth literacy also emerged as a significant determinant of satisfaction. Providers with higher literacy were more likely to be satisfied with the system. A possible explanation is that greater digital competence enables users to navigate EMRs more efficiently, interpret outputs accurately, and integrate digital information into clinical decision-making, thereby enhancing satisfaction. This finding is consistent with the eHealth Literacy Model and evidence from other scholars, where eHealth literacy was identified as a driver to IS success (31,47,48).

However, contrasting findings have been reported. Dubale et al. observed that eHealth literacy lost its independent association with satisfaction in the multivariable model. This discrepancy may reflect contextual or methodological differences, including variability in digital infrastructure, the scope of training interventions, or differences in how eHealth literacy was measured across studies.

It is equally important to note that not all user-related variables showed predictive value. For instance, LHIMS training did not emerge as a determinant of satisfaction. This evidence is consistent with the findings of Alasmary and Househ et al.(49) and Walle et al. (48), who similarly reported no significant relationship between basic technological training and satisfaction. In contrast, several other empirical studies, as well as established IT adoption frameworks such as the Technology Acceptance Model (TAM), have documented a positive and often strong association between training and satisfaction(5,10,13,50).

A plausible explanation for this apparent inconsistency lies in the training profile of the study population. Nearly all providers surveyed (94%) had already received training, leaving minimal variability for statistical discrimination. In such circumstances, where training coverage approaches universality, a ceiling effect occurs, thereby diminishing the explanatory power of training as a differentiating factor for satisfaction.

Similarly, computer literacy was not a determinant of user satisfaction, despite nearly half of the providers demonstrating good literacy levels. This contrasts with findings from studies conducted in Ethiopia, Saudi Arabia, and Australia, where computer literacy has been strongly associated with EMR satisfaction (5,33,48). A possible explanation is that the measure of literacy used may not have fully captured the operational skills required for effective system use. Field observations revealed that many providers had limited typing skills, which hindered documentation and data entry despite general familiarity with computers. Because these limitations were widespread, they did not translate into measurable differences in satisfaction. Overall, our findings suggest that satisfaction with LHIMS depends less on foundational skills such as basic computer literacy or universal training and more on the combination of enabling resources and advanced digital competencies. This highlights the need for capacity-building initiatives that move beyond basic training to develop advanced digital skills for sustained use of LHIMS.

Besides user background, several system-related constructs also influenced satisfaction. System quality was a significant predictor, as providers who perceived LHIMS to be stable, responsive, and user-friendly reported higher satisfaction. This finding resonates with evidence from Tanzania and Greece(51,52). A possible explanation is that reliable systems inspire user confidence, reduce workflow interruptions, and enable providers to focus on patient care rather than troubleshooting technical issues.

System usability was also a determinant. Providers who considered LHIMS simple, consistent, and easy to navigate were more satisfied, a pattern consistent with studies in Ethiopia (5,10). Usable systems reduce the cognitive burden on providers, shorten the learning curve, and fit more smoothly into clinical routines. Conversely, challenges such as navigating multiple tabs to retrieve patient information increase workload and frustration, diminishing satisfaction. These findings highlight the importance of designing user-friendly interfaces that align with workflow realities.

Information quality similarly played a critical role. Providers who perceived LHIMS outputs as precise, accurate, and valid reported higher satisfaction. This observation is consistent with studies in Ethiopia, Nigeria and Brazil, which have shown a strong relationship between EMR satisfaction and the quality of information produced(5,48,53). The DeLone and McLean IS Success Model also supports this, positing that good information quality is essential to system success(54). One possible explanation is that electronic systems produce structured and legible documentation, unlike handwritten records, which are often illegible and prone to errors. Clearer outputs improve communication among providers, reduce duplication of work, and enhance clinical decision-making, thereby increasing users’ satisfaction.

However, not all system-related factors were significant. Service quality, despite being a central construct of the DeLone and McLean Information Systems Success Model, did not predict satisfaction in this study. Similar findings have been reported in Ethiopia and Rwanda(41,48). By contrast, evidence from higher-resource contexts such as Canada, Saudi Arabia, and Brazil indicated that service quality characterised by responsive technical support was a strong determinant of EMR satisfaction (55–57). This contrast suggests that in resource-limited settings, service quality may not exert the same influence as in higher-resource health systems.

A plausible explanation is that broader infrastructural constraints common in low-resource settings, including unstable electricity, poor internet connectivity, and prolonged downtime, tend to overshadow perceptions of service quality. In Ghana specifically, these systemic challenges were compounded by concerns from health leaders that providers were not adequately consulted during the design and implementation of LHIMS, with their inputs not duly incorporated (21). These contextual issues may therefore reduce the explanatory power of service quality within the D&M model, as structural barriers and limited participatory design weaken its potential impact on user satisfaction.

Overall, system-related attributes, particularly quality, usability, and information outputs, proved central to provider satisfaction with LHIMS. These findings highlight the importance of optimising system performance, ensuring usability, and safeguarding information standards to strengthen user confidence.

## Conclusion

This study found that healthcare providers’ satisfaction with the Lightwave Health Information Management System (LHIMS) in Ghana was moderate. Satisfaction was relatively high for system speed, efficiency, and information quality, but low regarding the system’s impact on patient safety and quality of care. Significant determinants of satisfaction included user-related factors such as computer access and eHealth literacy, alongside system-related attributes including system quality, usability, and information quality. In contrast, LHIMS training, baseline computer literacy, and service quality were not significant, likely due to contextual barriers. The findings also highlight that limited end-user consultation during system design contributed to usability challenges and unmet workflow needs.

To enhance satisfaction, attention should be directed toward strengthening ICT infrastructure by ensuring reliable electricity and internet connectivity, while also improving equitable access to computers across facilities. Continuous investment in eHealth literacy is essential, not only through initial training but also through refresher and professional development programs. Incorporating provider feedback into system upgrades will help address usability limitations and better align LHIMS with clinical workflows. Furthermore, improving the accuracy, clarity, and security of system outputs will be crucial to establishing user trust and enhancing the system’s contribution to patient safety and quality of care.

## Limitations and Strengths

This study has certain limitations. First, the reliance on self-reported data may have introduced social desirability bias. Future studies could mitigate this by triangulating self-reported satisfaction with objective usage metrics such as system log files, error rates, or task completion times. The use of a cross-sectional design restricts causal inference and does not allow assessment of changes in satisfaction over time. Longitudinal studies would be valuable in capturing trends as LHIMS matures, while qualitative studies could complement survey findings by providing deeper insights into contextual challenges and user experiences.

Despite these limitations, the study has notable strengths. The use of validated, theory-driven instruments grounded in the DeLone and McLean Information Systems Success Model enhanced the credibility and theoretical rigour of the findings. Moreover, the inclusion of facilities across primary, district, and regional levels, combined with a multistage sampling approach using stratified proportional allocation and an adjusted design effect, improved representativeness, minimised clustering bias, and ensured methodological rigour. These methodological strengths, combined with the diversity of study settings, increase the generalizability of the findings and provide reliable evidence to inform digital health policy and practice in Ghana.

## Data Availability

All relevant data are within the manuscript and its Supporting Information files.

## Acknowledgement

We are grateful to the survey participants for their participation and valuable feedback. We also thank the management, human resources department, and health information department of the selected healthcare facilities for their invaluable support during the study. We also extend our sincere gratitude to the Trainer of Trainees of Lightwave e-Health Solutions and the research assistants for their contributions to this study.

## Authors’ contributions

Conceptualisation and design: Anane Bernard Gyamfi and Victoria Bam Data Curation: Anane Bernard Gyamfi, Gertrude Boafo Analysis and Interpretation: Anane Bernard Gyamfi and Osei-Peprah Emmanuel Drafting of Manuscript: Anane Bernard Gyamfi and Jacob Tetteh Revision of Manuscript: Victoria Bam

## Supporting Information

S1 Text: Data collection instrument

